# A blended learning approach for capacity strengthening to improve the quality of integrated HIV, TB, and Malaria services during antenatal and postnatal care in LMICs: A feasibility study

**DOI:** 10.1101/2023.05.04.23289508

**Authors:** Alice Norah Ladur, Elizabeth Adjoa Kumah, Uzochukwu Egere, Florence Mgawadere, Christopher Murray, Marion Ravit, Sarah Ann White, Hauwa Mohammed, Rael Mutai, Lucy Nyaga, Duncan Shikuku, Issak Bashir, Olubunmi Olufunmilola Ayinde, Rukia Bakar, Leonard Katalambula, Carlo Federici, Aleksandra Torbica, Nicholas Furtado, Charles Ameh

## Abstract

The blended learning (BL) approach to training health care professionals is increasingly adopted in many countries because of high costs and disruption to service delivery in the light of severe human resource shortage in low resource settings. The Covid-19 pandemic increased the urgency to identify alternatives to traditional face-to-face (f2f) education approach. A four-day f2f antenatal care (ANC) and postnatal care (PNC) continuous professional development course (CPD) was repackaged into a 3-part BL course; 1) self-directed learning (16 hours) 2) facilitated virtual sessions (2.5 hours over 3 days) and 3) 2-day f2f sessions. This study assessed the feasibility, change in healthcare providers’ knowledge and costs of the BL package in Nigeria, Tanzania, and Kenya. A mixed methods design was used. A total of 89 healthcare professionals, were purposively selected. Quantitative data was collected through an online questionnaire and skills assessments, analyzed using STATA 12 software. Qualitative data was collected through key informant interviews and focus group discussions, analysed using thematic analysis.

Majority of participants (86%) accessed the online sessions using a mobile phone from home and health facilities. The median (IQR) time of completing the self-directed component was 16 hours, IQR (8, 30). A multi-disciplinary team comprising of 42% nurse-midwives, 28% doctors, 20% clinical officers and 10% other healthcare professionals completed the BL course. Participants liked the BL approach due to its flexibility in learning, highly educative/relevant content, mixing of health worker cadres and CPD points. Aspects that were noted as challenging were related to personal log-in details and network connectivity issues during the self-directed learning and facilitated virtual sessions respectively. The blended learning approach to ANC-PNC in-service training was found to be feasible, cost saving compared to the face-to-face approach and acceptable to health care professionals in LMICs.

## Introduction

Globally, maternal mortality ratio declined from 339 maternal deaths per 100,000 live births in 2000 to 223 in 2020, a 34.3% reduction over the 20-year period (1). However, this far exceeds the global Sustainable Development Goal target of less than 70 per 100,000 (2). Although slight reduction in global MMR has been reported recently, sub-Saharan Africa still has the highest burden of the maternal mortality (545 maternal deaths per 100,000 live births) accounting for about 70% of maternal deaths in 2020 alone (1). Similarly, there has been an increase in births attended by skilled health personnel globally, but only 68% in low income and 78% in lower-middle-income countries are assisted by such skilled health personnel (3).

Most of the maternal deaths result from complications during and following pregnancy are preventable or treatable. Direct obstetric causes including haemorrhage before and after childbirth, hypertensive disorders, infections/sepsis after childbirth, complications from delivery and abortion account for about 75% of the maternal deaths globally (4). Global evidence shows that HIV, malaria, and TB-related complications are among the leading causes of indirect maternal mortalities and morbidities (4). In Kenya, the national confidential enquiry into maternal deaths in 2017 showed that HIV-related complications (22.9%) and malaria (10.4%) were among the leading indirect causes of maternal mortality (5). To reduce the maternal deaths, WHO through the Ending Preventable Maternal Mortality (EPMM) strategy recommends that countries should focus on addressing inequalities in access to and quality of reproductive, maternal, and newborn health care services; ensuring universal health coverage for comprehensive reproductive, maternal, and newborn health care; and addressing all causes of maternal mortality, reproductive and maternal morbidities, and related disabilities (6).

Building capacity of healthcare professionals through “in-house” trainings facilitates the acquisition of knowledge, skills, and competence to provide integrated antenatal (ANC) and postnatal care (PNC) services, contributing to quality of care in maternity services (7). Regular training is recommended to ensure continued accreditation of healthcare professionals (8).

A blended learning (BL) approach to training health care professionals is increasingly adopted in many countries to reduce prohibitive costs and disruption to service delivery in the light of severe human resource shortages in low resource settings (9). Blended learning presents a unique approach to offer “in-service or “on-the job training” for healthcare professionals involved in the provision of integrated antenatal and postnatal care services. Evidence suggests that a blended learning approach is as effective as conventional approaches in increasing providers’ knowledge while reducing costs (9, 10). Lessons learnt from the COVID-19 pandemic, highlighted the need to identify alternatives to traditional face-to-face training approaches in pre-service midwifery education in Kenya (11, 12). We developed and piloted a blended learning training package, in Kenya, Nigeria, and Tanzania, for healthcare providers in integrated TB, HIV and Malaria services in ANC and PNC as an alternative to our previously used face-to-face training package. This study’s objectives were to determine the feasibility, acceptability, change in healthcare providers’ knowledge and describe the cost of developing and implementing the blended learning package.

## Materials and Methods

We conducted a feasibility and acceptability study of an ANC-PNC blended learning training course in Nigeria, Tanzania, and Kenya. The specific objectives of the study were to 1) explore the acceptability and feasibility of implementing our ANC and PNC blended learning course for healthcare providers in Nigeria, Tanzania, and Kenya, 2) assess the change in knowledge of healthcare providers after implementation of the ANC and PNC blended learning course and 3) to describe the cost of developing the package, and compare the cost of its implementation in each of the study countries.

### Description of the ANC-PNC blended learning training course

In collaboration with international stakeholders a multi-disciplinary team from Liverpool School of Tropical Medicine (LSTM) developed the ANC-PNC blended learning training course, using WHO recommendations and guidelines in the study countries (13). The course is designed to strengthen the capacity of healthcare professionals to provide quality integrated HIV, TB and Malaria Services in Antenatal care and Postnatal care in LMICs. It covers essential components of ANC and PNC, such as quality care, respectful maternity care, communication, mental health assessment and integration of HIV, TB, and malaria services. The target audience are nurses, midwives, clinical officers, medical assistants, doctors, and other cadres of health care providers involved in the provision of ANC-PNC in LMICs.

The blended learning training was completed in three parts: **1) self-directed learning**. Course instructions, pre-recorded lectures and reading materials were uploaded onto an online platform developed by the World Continuing Education Alliance (WCEA). Health professionals engaged with online course content at their own pace and timing over a period of two weeks (16 hours or 39% of the whole course) in August 2022. Upon completion of the self-directed learning, they are invited to complete part two: **2) virtual facilitated sessions**. In the second BL component, health professionals attended facilitated live sessions delivered via zoom over three days, lasting 2.5 hours per day (7.5 hours in total, or 18% of the whole course) in August 2022. Upon completion of the two BL components, learners were invited to complete the last part of the blended learning course: **3) face-to-face sessions**. Learners and faculty attended this 2-day component in-person (18 hours or 43% of the whole course) in October 2022. During this component, health professionals engaged in activities designed to enable them to acquire hands-on skills and apply the theoretical concepts covered in the self-directed and virtual facilitated components. The component was delivered by experienced LSTM in-country staff/implementation partners based in that country and Ministry of Health in the three countries.

### Study design

Feasibility studies may be used to determine whether an intervention can be carried out and the appropriateness of the intervention from the recipient’s perspective (14). This study used a mixed-methods design with quantitative and qualitative data collected concurrently, analyses done independently and mixed/integrated during the overall discussion of findings (15).

The Kirkpatrick’s (1996) model was adopted to evaluate the feasibility and acceptability of the ANC-PNC blended learning training course. Kirkpatrick’s model consists of four levels of evaluation of a training programme: 1) reaction: this determines how learners engaged with the training and how actively they contributed to the training (including acceptability of the training programme); 2) learning: this assesses the learner’s learning outcomes and improvement in knowledge, skills, and attitudes towards the training experience (16). This level is usually measured using a pre- and post-test assessment questionnaire (La Duke, 2017); 3) behaviour, which determines whether the training resulted in improvement in practice at the workplace; and 4) results: this assesses the overall impact of the training programme based on the programme objectives and it determines the cost-effectiveness of the training programme, improved quality, client satisfaction, as well as improved productivity. The study assessed only two levels of the Kirkpatrick model: 1) level 1 (acceptability and feasibility of the ANC-PNC blended learning approach) 2) level 2 (change in knowledge). A third objective beyond the scope of the Kirkpatrick model was added to 3) describe the cost of developing and implementing the blended learning training and the implementation costs was compared with that for the face-to-face only approach.

### Recruitment and sampling

A total of 89 healthcare professionals from Nigeria, Kenya, and Tanzania, were purposively selected from health care facilities agreed with the Ministry of Health in each country to participate in the ANC-PNC blended learning course and feasibility study. The sample of 89 participants falls within the recommended sample size of at least 70 for a pilot or feasibility study, to reduce the imprecision around the estimate of the standard deviation (17). The criteria for inclusion in the feasibility study consisted of health facilities in the implementation program areas, healthcare professionals involved in the provision of antenatal or postnatal care services in the identified health facilities and consent for taking part in the blended learning and/or data gathering activities. The cadres selected consisted of nurses, midwives, clinical officers, medical officers, and community health workers from rural and urban health facilities (Table 1). The exclusion criteria involved healthcare professionals who did not undertake the ANC-PNC blended learning course and anyone not providing consent to take part in the study.

**Table 1:**
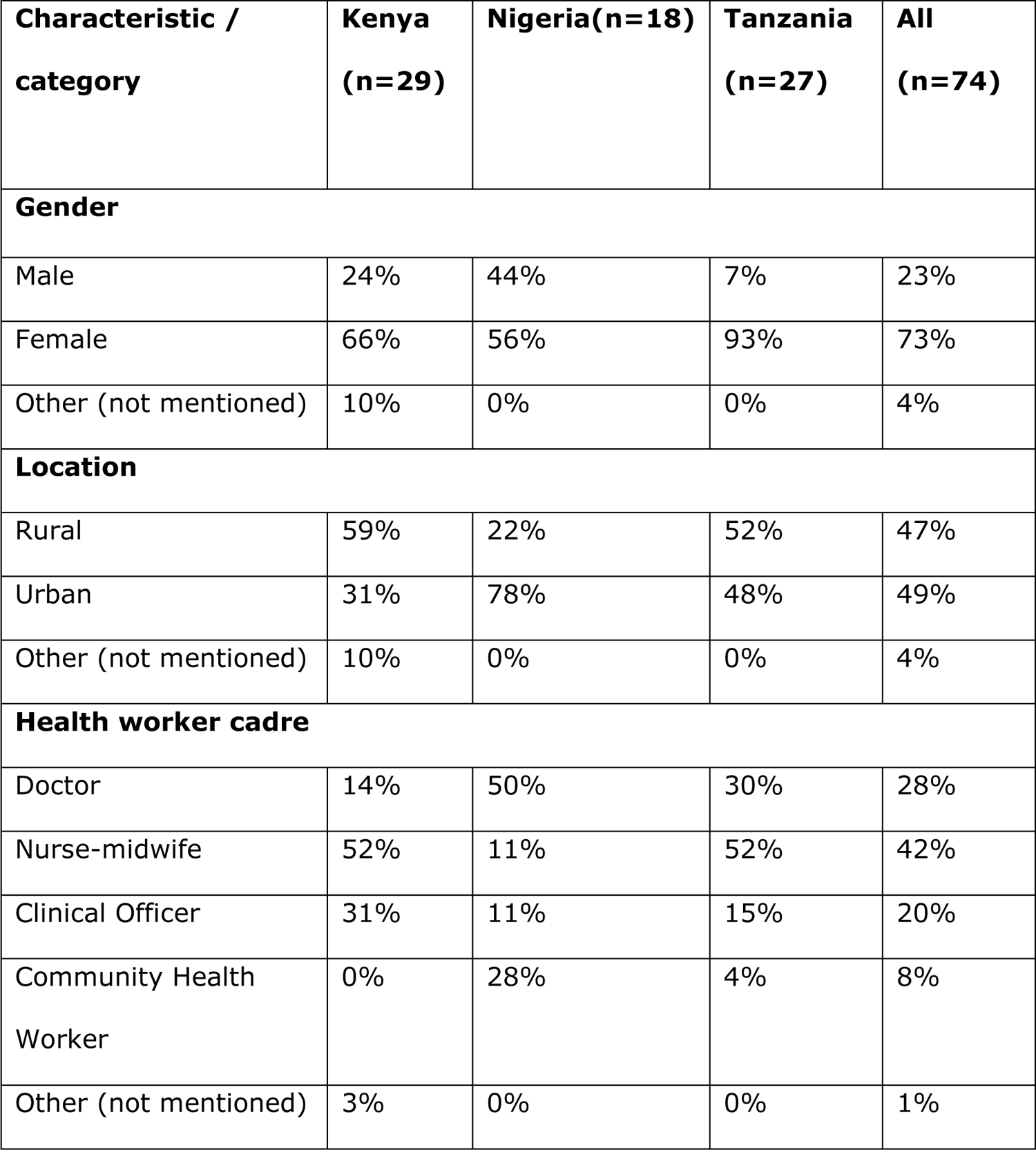
Characteristics of participants by country

| Characteristic / category | Kenya (n=29) | Nigeria(n=18) | Tanzania (n=27) | All (n=74) |
| --- | --- | --- | --- | --- |
| **Gender** |  |  |  |  |
| Male | 24% | 44% | 7% | 23% |
| Female | 66% | 56% | 93% | 73% |
| Other (not mentioned) | 10% | 0% | 0% | 4% |
| **Location** |  |  |  |  |
| Rural | 59% | 22% | 52% | 47% |
| Urban | 31% | 78% | 48% | 49% |
| Other (not mentioned) | 10% | 0% | 0% | 4% |
| **Health worker cadre** |  |  |  |  |
| Doctor | 14% | 50% | 30% | 28% |
| Nurse-midwife | 52% | 11% | 52% | 42% |
| Clinical Officer | 31% | 11% | 15% | 20% |
| Community Health Worker | 0% | 28% | 4% | 8% |
| Other (not mentioned) | 3% | 0% | 0% | 1% |

### Qualitative methods and analysis

Eight focus group discussions (FGDs) and nine key informant interviews (KIIs) were held after the ANC-PNC BL course in October 2022. KIIs lasted for thirty minutes and FGDs for 60 minutes. In Nigeria, two focus group discussions were held with nurse-midwives and community health workers separately to allow the participants to freely share their views. In Kenya and Tanzania three FGDs were held in each country. The FGDs were mixed as per the request of the health workers who chose to stay in their learning groups whilst sharing experiences of the blended learning approach. Three key informant interviews were held with participants from each of the study sites and considered adequate after obtaining saturation. A semi-structured topic guide was used to guide discussions in the KIIs and FGDs. All interviews were audio recorded with permission from the participants and transcribed verbatim into English. KIIs and FGDs were helpful in eliciting information on perceptions and experiences of the blended learning approach and ANC-PNC course content (18).

Preparation for qualitative data analysis involved a rigorous process of transcription, data reduction (coding) and theme development (19). Qualitative data was analysed in NVIVI-O 12 software using inductive thematic analysis by Braun and Clarke (2006); a) familiarising oneself with the data through transcription and reading transcripts; b) generating initial codes; c) searching for themes; d) reviewing themes; e) defining and naming themes; and g) writing findings/producing a report. Pseudonyms were used to maintain confidentiality in the study. Trustworthiness was achieved by using a criterion for thematic analysis; returning to the data repeatedly to check for accuracy in interpretation; discussions with the study team with expertise in midwifery, blended learning, and mixed methods research (18).

### Quantitative methods and analysis

Quantitative data was collected in two phases, 1) Knowledge through an online questionnaire in August and, 2) skills assessments in October 2022 after the final part of the BL course. For the face-to-face component, each participant was asked to complete a knowledge assessment (MCQs) and two OSCEs (breastfeeding and insertion of an IUD) before starting training and immediately after training. The same tools were used by all participants on both occasions. All questionnaires were completed anonymously. Changes in percentage scores are constrained by the pre-training score, with those scoring well having less capacity for improvement than those scoring poorly. To account for this in analysis, the improvement percentage score (IPS) was derived as follows:

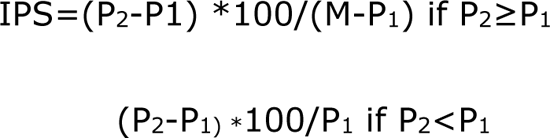

where: P_1_ denotes the pre-training percentage score; P_2_ denotes the post-training percentage score, and M denotes the maximum possible score (100 for knowledge and 20 for OSCEs). For each type of assessment, analysis of covariance (ANCOVA) was then used to analyse the improvement percentage scores, with pre-training scores as the covariate and factors for each country. The 5% significance level was used to determine statistical significance. Means are reported with 95% confidence intervals.

### Cost analysis

The cost analysis considered both the design costs of the BL training package as well as the cost of implementation in each country. For Tanzania, costs were reported separately for the BL training implemented in Dodoma and Zanzibar. To calculate the full economic cost of the BL training packages, both the perspectives of the designer and implementers of the training and the costs of the recipients of the course were considered. For the recipients of the BL training, the labor cost of HCPs participating in the training was calculated by multiplying the time spent overall to attend the different components of the training package times the average salary of their role and level. For the costs of the implementer of the training, economic costs for both the design and the implementation of the BL training package were reported, grouped by resource categories which included direct and indirect costs. Direct costs included costs for direct labor, per-diems and travel allowances, costs for face-to-face events (e.g., workshop venues, catering), transport costs, and printing of training materials. For the implementation in each country indirect costs such as indirect labour, use of capital and overheads were assumed to be 7% of the total direct costs.

Costs were reported as the total cost of design of the BL training and the cost of implementation in each country. For the latter, the total cost per participant was also calculated. All costs were converted from local currencies to 2021 USD. Conversion factors were taken from the World Bank and were equal to 0.73 for GBPs, 109.64 for Kenyan Shillings, 2,297.76 for Tanzania Shillings and 358.81 for Nigerian Naira.

## Results

### Quantitative findings

#### Characteristics of participants and access to training platform

Of the 89 participants selected for the study, only 74 (83%) completed the self-directed learning component of the blended learning course whilst 80 participants completed the facilitated virtual component (delivered via zoom), and 89 health workers completed the face-to-face component.

The demographic characteristics of the participants in the self-directed learning are shown in table 1. Overall, a majority (73%) of the 74 participants were females and 49% were providing healthcare in urban settings. The cadre of participants included nurse midwives (42%), doctors (28%) and clinical officers (20%).

The average time to completion of the self-directed learning component was 16 hours, IQR (8, 30). The participant owned mobile phone was the most used device (86% vs. 14%) compared to other devices. Most of the participants (58%) joined the live zoom sessions from the health facilities whilst 27% joined from their homes.

### Outcome of knowledge and skills assessment

#### Outcome of knowledge

Data were received for 72 trainees with pre and post test scores: 29 from Kenya, 21 from Nigeria, 28 from Tanzania. Two trainees did not have post-test scores and were therefore excluded from the analysis. Data for knowledge assessment on one occasion was not received for two participants in Kenya (post-training), three participants in Nigeria (pre-training) and 1 participant in Tanzania (pre-training). Data for all OSCEs was missing for 5 participants in Nigeria and on one occasion for the Breast feeding OSCE for Tanzania. Knowledge was assessed in 72 participants both before and after training. In each country, the mean knowledge score before training was between 60% and 70% (Table 2) and the individual scores ranged between 40% and 87%. Overall, the mean (95% CI) absolute improvement in scores was 10% (7%,13%) and the mean (95% CI) percentage improvement was 27% (21%,34%); on both scales, the mean improvements observed for Kenya and Nigeria were larger than in Tanzania. ANCOVA of the percentage improvement scores found statistically significant evidence that the higher the baseline score the smaller the percentage improvement score was (p=0.001, Table 3) and that there were significant differences between countries (p=0.003). For Tanzania, the percentage improvement was statistically significantly smaller than for Kenya (p=0.002) whereas there was no significant difference between Nigeria and Kenya.

**Table 2:**
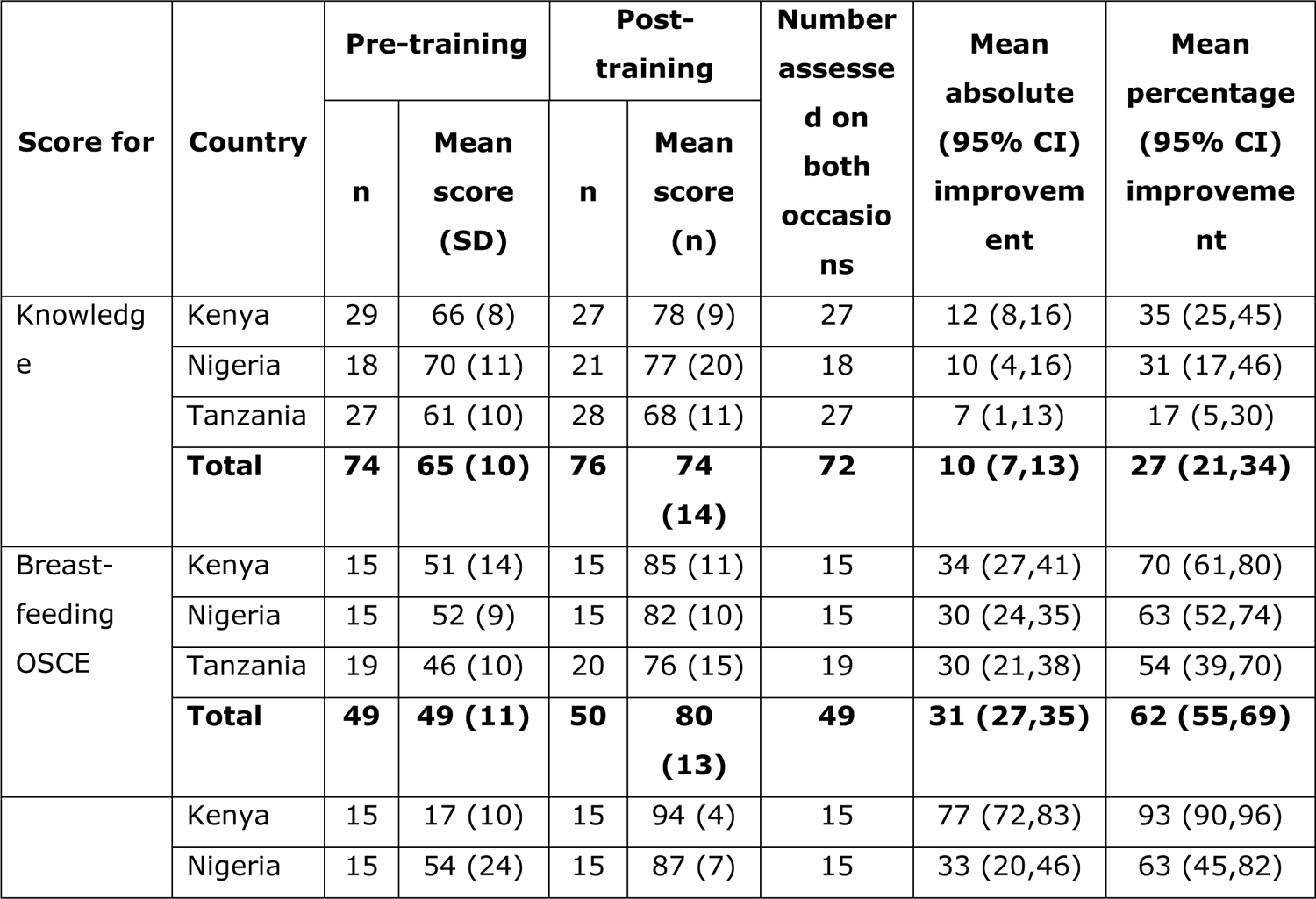
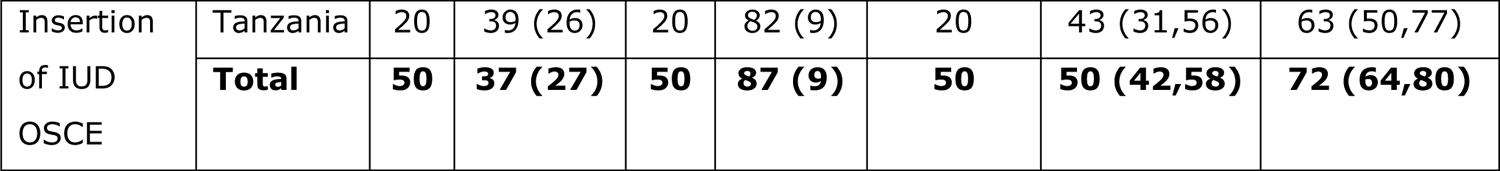
Summary of changes in scores, by country

**Table 3:** Estimates from analyses of covariance of improvement percentage scores

|  | Knowledge |  | Breastfeeding OSCE |  | Insertion of IUD OSCE |  |
| --- | --- | --- | --- | --- | --- | --- |
|  | Estimate<br>(95% CI) | p-value | Estimate<br>(95% CI) | p-value | Estimate<br>(95% CI) | p-value |
| **Constant** | 114.2<br>(69.4,159.0) |  | 67.0<br>(30.8,103) |  | 104.6<br>(92.8,116) |  |
| **Pre-**<br>**training**<br>**coefficient** | -1.20<br>(-1.87, -0.54) | 0.001 | 0.07<br>(-0.60,0.73) | 0.84 | -0.71<br>(-0.98, -0.44) | <0.001 |
| **Variation**<br>**between**<br>**countries** |  | 0.003 |  | 0.21 |  | 0.13 |
| **Nigeria v**<br>**Kenya** | 1.66<br>(-14.8,18.1) | 0.84 | -7.4<br>(-25.5,10.7) | 0.42 | -2.7<br>(-21.1,15.8) | 0.77 |
| **Tanzania v**<br>**Kenya** | -23.7<br>(-38.5, -8.9) | 0.002 | -15.6<br>(-33.0,1.9) | 0.08 | -13.8<br>(-29.3,1.7) | 0.08 |

#### Outcome skills assessment

The breastfeeding OSCE was completed by 49 participants both before and after training. In each country, the mean percentage score before training was between 46% and 52% and individual OSCE scores were between 30% and 75% (Table 3). Overall, the mean (95% CI) absolute improvement in scores was 31% (27%,35%) and the mean (95% CI) percentage improvement score was 62% (55%,69%). ANCOVA did not find evidence that the pre-training score had any effect on the percentage improvement scores (p=0.84, Table 3). No statistically significant differences were detected between the countries (p=0.21).

The insertion of IUD OSCE was completed by 50 participants both before and after training. There were substantial differences in score between country before training, with the mean (SD) score for Kenya being 17% (10%) whereas for Nigeria, the mean was 54% (24%) (Table 2). Individual percentage scores ranged between 0% and 95%. After training, the mean scores for the three countries were more similar, ranging between 82% and 94%. Overall, the mean (95% CI) absolute improvement in scores was 50% (42%,58%) and the mean (95% CI) percentage improvement was 72% (64%,80%). ANCOVA of the percentage improvement scores found statistically significant evidence that the higher the baseline score the smaller the percentage improvement score was (p<0.001, Table 3), but there were no significant differences between the three countries (p=0.13).

### Cost of designing and implementing the BL training approach

The cost of designing the BL training package was estimated at USD 22,749 and was mostly determined by labor costs for designing the BL format (200 person hours in total) and coordinating meetings for subsequent implementation in each country (Approximately 24 person hours in total for the 4 implementation sites) (Table 4).

**Table 4:** Costs of designing the blended learning training package (USD)

|  | Quantity | Unit cost | Total cost |
| --- | --- | --- | --- |
| **Design of BL training package** |  |  |  |
| **Labor costs** |  |  |  |
| Packaging of course content into BL format (person hours) | 200 | 42 | 8,400 |
| Recording of lectures (person hours) | 20 | 52 | 1,040 |
| Planning meetings for roll out of BL course (person hours) | 96 | 40 | 3,853 |
| **Equipment** |  |  |  |
| Direct WCEA platform design costs for SDL component (units) | 1 | 7,968 | 7,968 |
| **Overheads (7% of direct costs)** |  |  | 1,488 |
| **Total** |  |  | **22,749** |

Roll out costs were equal to USD 15,773; 13,525; 25,804 and 25,152 respectively in Tanzania-Dodoma, Tanzania-Zanzibar, Kenya, and Nigeria (Table 5). Most of this cost was incurred by the implementers (Direct costs) of the BL training, accounting for 90.10%, 92.16%, 71.74% and 80.18% of total costs respectively in the four implementation sites, whereas the reminder cost was related to the time spent by participants to attend the course. With the only exception of Zanzibar, the total cost per participant was similar in the other implementation sites and ranged between USD 830 in Dodoma and USD 860 in Kenya when considering both costs of implementers and participants, or between USD 624 and USD 759 when considering only the costs to the implementers. In Zanzibar, the cost per participant was higher, at USD 1,353 (USD 1,267 when considering only direct costs (the costs to the implementers), due to the smaller number of participants and the higher incidence of fixed costs.

**Table 5:**
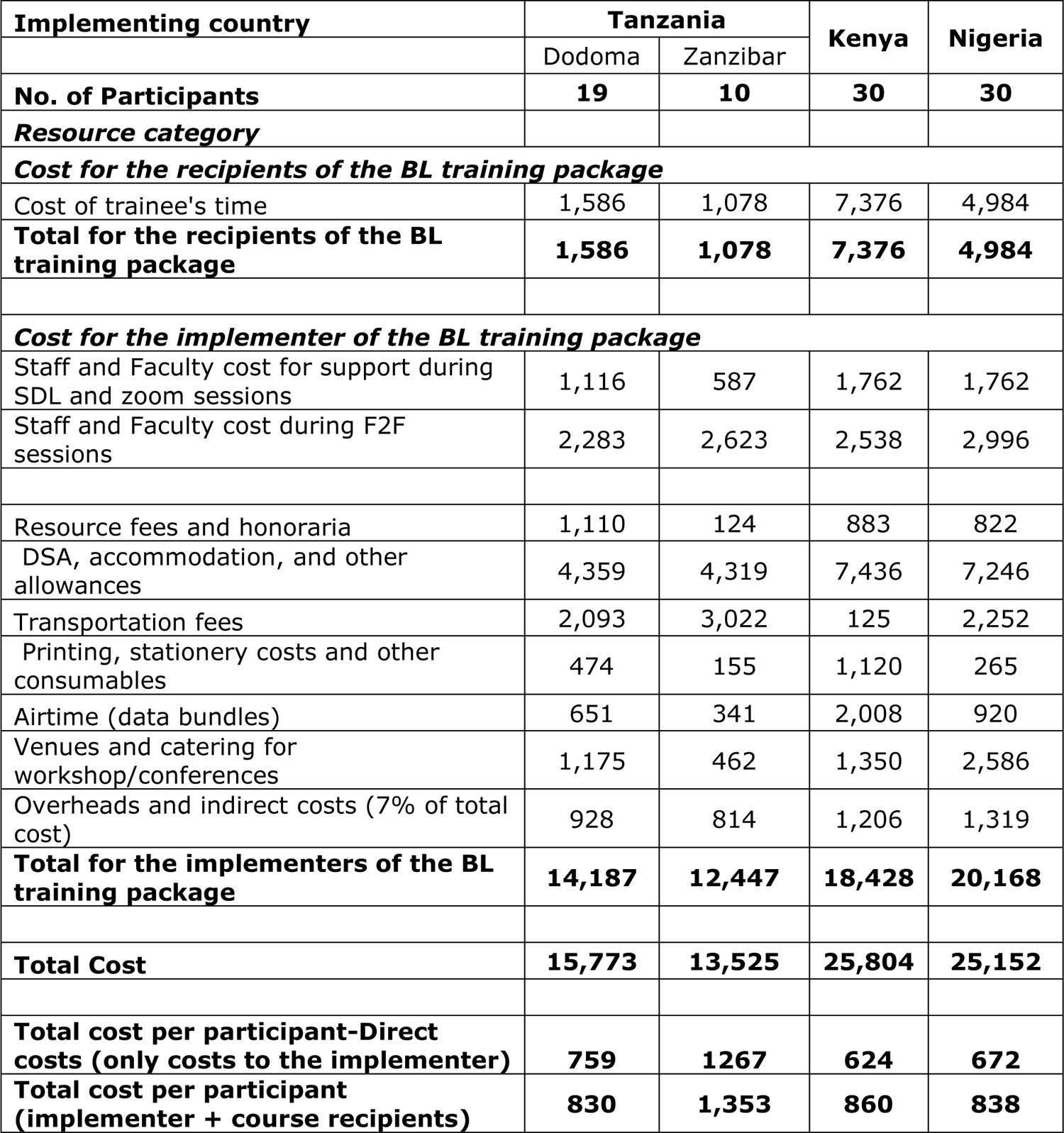
Costs of rolling out the BL training package (USD)

| Implementing country | Tanzania |  | Kenya | Nigeria |
| --- | --- | --- | --- | --- |
|  | Dodoma | Zanzibar |  |  |
| **No. of Participants** | **19** | **10** | **30** | **30** |
| **Resource category** |  |  |  |  |
| ***Cost for the recipients of the BL training package*** |  |  |  |  |
| Cost of trainee's time | 1,586 | 1,078 | 7,376 | 4,984 |
| **Total for the recipients of the BL training package** | **1,586** | **1,078** | **7,376** | **4,984** |
| ***Cost for the implementer of the BL training package*** |  |  |  |  |
| Staff and Faculty cost for support during SDL and zoom sessions | 1,116 | 587 | 1,762 | 1,762 |
| Staff and Faculty cost during F2F sessions | 2,283 | 2,623 | 2,538 | 2,996 |
| Resource fees and honoraria | 1,110 | 124 | 883 | 822 |
| DSA, accommodation, and other allowances | 4,359 | 4,319 | 7,436 | 7,246 |
| Transportation fees | 2,093 | 3,022 | 125 | 2,252 |
| Printing, stationery costs and other consumables | 474 | 155 | 1,120 | 265 |
| Airtime (data bundles) | 651 | 341 | 2,008 | 920 |
| Venues and catering for workshop/conferences | 1,175 | 462 | 1,350 | 2,586 |
| Overheads and indirect costs (7% of total cost) | 928 | 814 | 1,206 | 1,319 |
| **Total for the implementers of the BL training package** | **14,187** | **12,447** | **18,428** | **20,168** |
| **Total Cost** | **15,773** | **13,525** | **25,804** | **25,152** |
| **Total cost per participant-Direct costs (only costs to the implementer)** | **759** | **1267** | **624** | **672** |
| **Total cost per participant (implementer + course recipients)** | **830** | **1,353** | **860** | **838** |

Of the total costs to the implementer (Direct costs), costs for DSA, accommodation and other allowances were the major cost driver accounting for respectively 30% in Dodoma, 34% in Zanzibar, 40% in Kenya and 36% in Nigeria. Labour costs were the second highest cost driver in all implementation sites, accounting in a range from 23% to 25% of the total cost in the four implementation sites. Other relevant costs were transportation fees in Tanzania (respectively 15% and 24% of the total cost in Dodoma and Zanzibar) and Nigeria (11% of the total cost), the cost for venues and catering for workshop/conferences in Nigeria (13% of the total cost), and the cost for airtime data bundles in Kenya (11% of the total cost).

The F2F component of the training package accounted for 66.7% to 86.8% of the total costs to the direct costs in the four sites (Table 6).

**Table 6:** Cost by component of the BL training package (USD) and % incidence over total implementer cost

| Implementing country | Tanzania |  |  |  | Kenya |  | Nigeria |  |
| --- | --- | --- | --- | --- | --- | --- | --- | --- |
|  | Dodoma |  | Zanzibar |  | Nakuru |  | Oyo State -BL |  |
|  | Total cost (cost per participant) | % total BL costs | Total cost (cost per participant) | % total BL costs | Total cost (cost per participant) | % total BL costs | Total cost (cost per participant) | % total BL costs |
| **F2F** | 10,938 (576) | 82.5 % | 10,102 (1010) | 86.8 % | 11,485 (383) | 66.7 % | 14,710 (490) | 78.0 % |
| **SDL** | 1,492 (79) | 11.2 % | 889 (89) | 7.6 % | 3,362 (112) | 19.5 % | 2,236 (75) | 11.9 % |
| **Virtual Learning** | 830 (44) | 6.3 % | 642 (64) | 5.5 % | 2,375 (79) | 13.8 % | 1,902 (63) | 10.1 % |
| **Total BL costs** | 13,259 (698) | 100.0 % | 11,633 (1163) | 100.0 % | 17,222 (574) | 100.0 % | 18,849 (628) | 100.0 % |

### Qualitative findings

The analysis of the qualitative data generated three themes about the BLA: 1) general perceptions on the approach; 2) challenges participating; 3) recommendations improving the BLA.

#### 1) Perceptions on the blended learning training approach Inclusive learning

All participants loved the blended learning approach to training in ANC-PNC across all the health worker cadres in Nigeria, Kenya, and Tanzania. It was a new training approach with diverse learning methods that caters to a variety of learners, as highlighted in this extract.

*“The blended course was a new and interesting learning process. Everyone is learning at every part of that training. Someone who is not good at reading on their own could complement the part in the zoom session. And those who could not comprehend the zoom session would complement on the face-to-face. We appreciate it because we are learning both on our own and being facilitated face to face. It was a good experience, and I liked it*.” **Clinical officer 2, Kenya**

Participants appreciated the three learning parts of the blended learning course which was inclusive and created opportunities for all learners to grasp the course content through self-study and facilitated sessions.

##### Alternative to classroom-based learning

Participants reflected on their experiences of learning during the ANC-PNC blended learning course as an eye opener to other modes of learning.

*The blended training is an eye opener that one can still learn even when not in the class. “I did not have issues with the network. So, all the lectures I was able to participate, some of them I would be driving and listening [audio recordings in self-directed learning]. It saved my time, and I was able to work. It is interesting and helpful*.” **Nurse midwife 1, Nigeria**

The ANC-PNC blended learning course is flexible, time saving, convenient and causes less disruption to work.

##### Blended learning sequence

Participants loved the learning sequence of theoretical concepts/knowledge preceding the face-to-face component. The online sessions provided an overview of the entire course highlighting topics which would be covered in each of the three parts of the blended learning course.

*“This program [training] is very nice because you come to face-to-face session when you know the content. Already you have read for yourself at home or at your workplace and have an idea of what you are going to do unlike other programs. That’s what was interesting about the program*.” **Doctor 1, Kenya**

Providing a course overview enabled the participants to know what to expect during the self-directed learning, zoom and face-to face components.

##### Educative course content

The course content was noted as highly educative, well-structured, and very helpful in addressing gaps in ANC-PNC service provision that would contribute to improving the quality of maternity care in the three countries.

“I want to really appreciate the organizers of this program because it was well planned, they really looked into the problem that we were facing as nurses and health workers and they tried to find solutions, like this respectful maternity care.” **Nurse midwife 4, Nigeria.**

During the zoom and face-to-face sessions, facilitators reinforced the need for health workers to provide respectful maternity care for women and families to improve on uptake of maternity services/health facility deliveries. Participants reflected on their health practice considering the knowledge and skills obtained from the course and mentioned several areas of improvement such as respectful maternity care, TB screening, documentation, referral, postnatal care, and mental health screening.

##### Target audience/mixing of health worker cadres

Consistent with the team approach to care provision, the ANC-PNC blended learning course included all health worker cadre providing ANC-PNC services.

“We also want to appreciate LSTM and the ministry [of health] for how they picked the participants across the cadres like the nurses, clinical officers, medical officers, which was a perfect one. Often with ANC (Antenatal Care) and PNC (Postnatal Care), it has been viewed as a nursing profession, but LSTM has done well to enlighten the other cadres and have everybody participating in the management of these mothers.” **Nurse 7, Kenya.** Participants were thankful for the unrestricted selection of learners across the health worker cadres which reinforced the need for teamwork in ANC-PNC service provision.

##### Mode of delivery

Participants reported that the Face-to-face sessions were most popular because they were hands-on to improve skills, allowed sharing of perspectives/experiences from different cadres, ensured maximum interaction with facilitators, and were conducted in an environment with no distractions that facilitated full concentration during the training.

“The phase which I understood the most and I got a lot of things was the face to face. For example, when I was reading through self-directed method, I was alone and there is no one to ask, but in face to face when I fail to understand something I just ask a teacher because they are present, also through this method I was practicing and sharing with my colleagues.” **Nurse midwife 9, Dodoma-Tanzania**

The self-directed learning component received mixed views with some participants suggesting it was worthwhile to engage in self-study at own pace and timing whilst others missed the interactive experience with peers and facilitators. Internet challenges and work commitments affected virtual learning making it the least liked component of the ANC-PNC blended learning course.

##### Planned behaviour

Participants shared topics that were intriguing and what they planned to do differently to improve on quality of maternity care in the health facilities where they worked in the extracts below.

“I realize we have not been doing good respectful maternity care. I will approach the health facility management committee so that we have a table, where we can receive our mothers, take proper history, because we have just been asking them through the window which is not good as there is no privacy” **Nurse midwife 5, Kenya**.

“I wish to improve on referral in our health facility as we do not have that referral form [SBAR (Situation Background Assessment Recommendation)]. What we have is not customized to SBAR as per guidelines. I would have to advocate to print the SBAR form and other documents including the whooleys and the Edinburgh [mental health screening tools],” **Clinical officer 10, Kenya**.

Participants reflected on their health practice considering the knowledge and skills obtained from the course and mentioned several areas of improvement such as respectful maternity care, referral & documentation systems, TB screening, postnatal care, and mental health screening.

#### 2) Challenges

Challenges experienced by participants in the three countries were similar despite different contexts and level of training such as the use of the WCEA online platform during the self-directed learning, use of zoom, internet connectivity, and work commitments during the virtual facilitated sessions.

##### Use of the WCEA platform

Participants experienced challenges with logging in to the WCEA platform to access the resources during the start of the self-directed learning. The log in challenges were related to incorrect passwords and how to navigate through the resources on the WCEA platform.

“The challenge I found is on the internet [self-directed learning] because I didn’t know how to log in or how to search for notes, but I am thankful to a member of the [WhatsApp] group. We sat together and she taught me until I understood, when I returned home, I could study by myself.” **Nurse midwife 6, Zanzibar-Tanzania**.

A WhatsApp group was created for participants in the three countries for communication purposes and to resolve challenges in real time. The WhatsApp groups also served as an interaction space for the participants to provide peer-to-peer support in completing the self-directed learning component of the course.

##### Work commitments

Zoom sessions were delivered in the afternoon for 2.5 hours in anticipation that health workers would use the morning hours to work in the ANC/PNC clinics, this was to minimise disruptions to service provision. The time was pragmatically adjusted in each context based on the clinic schedule.

“The challenge I faced is the same as my colleagues, zoom session time, most of time it starts at 1PM its time in which you may be handing over the report or there are many clients.” **Nurse midwife 3, Dodoma-Tanzania**.

“On the first day, I was still seeing patients about 45 minutes into the Zoom sessions. It would be very good if the zoom should happen between 5pm and 7pm after work”. **Doctor 1, Kenya.**

Initially, the scheduling of the zoom sessions coincided with busy times at the health facilities which became a challenge for some health workers to actively participate in the discussions, however, lessons were learnt from the experience and the schedule adjusted to minimise any disruptions to service delivery.

##### Internet connectivity issues

Poor internet posed a challenge for some participants joining the facilitated virtual sessions from remote areas and changes in weather patterns (torrential rain).

“During the zoom session, we have a change in the weather and the network starts fluctuating and misbehaving. I was lost during some of the lectures…” **Nurse midwife 7, Nigeria.**

Internet connectivity issues affected learning during the facilitated virtual sessions as participants would drop off from the Zoom sessions and try to reconnect once internet connection is restored.

##### Language barrier

Language barrier was a challenge unique to participants from Tanzania during the zoom sessions. Zoom sessions were facilitated in English, however, participants were more fluent in Swahili, a national language which limited interactions on day 1. The language issue was resolved on day two and three with facilitators using Swahili to facilitate the discussions. Lessons were learnt from the zoom experience and implemented in the face-to-face component of the blended learning course.

#### 3) Recommendations for the roll out of the ANC-PNC blended learning course

Participants were asked to share suggestions to improve the implementation of the ANC-PNC blended learning course. Most participants gave suggestions for all future in-house trainings in maternity services to be adapted to a blended learning format due to the learning techniques which were considered fun and supported active learning such as group discussions, breakout sessions and role plays.

The course delivery was wonderful because we were required to be active and participate fully in small groups. Other courses most of the time, have lectures which just make us sleepy. Other trainings should follow this type of method which we were using of blended learning. **Nurse 5, Dodoma-Tanzania**

Participants suggested a comprehensive induction to the blended learning course covering aspects such as use of zoom and WCEA platform. Notable in the recommendations were how to address the implementation gaps experienced in the virtual facilitated component of the blended learning course: 1) schedule zoom sessions in the evening/after work hours or over the weekend; 2) seek official permission and communication from health leaders to enable health workers participate; and 3) record live sessions to mitigate internet connectivity challenges.

## Discussion

This study sought to evaluate the feasibility of implementing the ANC-PNC blended learning course in low resource settings. An emphasis was placed on eliciting views on experiences/acceptability of the blended learning approach and identifying proactive strategies to improve future course implementation in LMICs settings. This study is unique because it used a flipped classroom (theoretical concepts preceded the practical skills) mode of the BL approach which enhanced active participation and learning to in-service training for frontline health professionals involved in the provision of ANC-PNC services in low resource settings (20).

### Acceptability/feasibility of the blended learning approach

The blended learning approach to training in ANC-PNC was found to be acceptable and feasible to implement in the study settings. A multi-disciplinary cohort of doctors, nurses/ nurse-midwives, clinical officers, and community health workers completed the blended learning course. Most participants accessed online sessions (self-directed learning and virtual facilitated sessions) using their phones from home or health facilities. Participants liked the blended learning approach due to its flexibility in learning, highly relevant/ educative content, less time spent away from work and link to CPD (Continuous Professional Development) points. Aspects that were noted as challenging were related to personal log-in details and network connectivity issues during the self-directed learning and virtual facilitated sessions, respectively.

### Knowledge and skills

Findings from this study indicate that the antenatal and postnatal care blended learning course was successful in improving the knowledge and skills of healthcare providers, immediately after the training compared to before. Participants reported improvements in knowledge on topics such as, respectful maternity care, referral systems, integration of HIV, TB, and Malaria services in ANC-PNC. In addition, participants mentioned the acquisition of new skills on postnatal IUCD insertion, breastfeeding, mental health/domestic violence screening. The knowledge and skills obtained from this course will be used to improve quality of integrated ANC-PNC care in health facilities where participants work. Results from this study are consistent with other studies reporting on the effectiveness of blended learning in enhancing knowledge acquisition among health professionals (21, 22). A study by Ameh et al 2019 reported that short competency-based trainings in maternity care led to significant improvements in health worker competence and changes in clinical practice (4).

### Costs

The cost of the BL learning package to the implementer in the four implementation sites were similar in Dodoma region of Tanzania, Kenya, and Nigeria, and ranged between USD 624 and USD 759. In Zanzibar, the cost per participant was higher mostly due to the small number of participants which increased the share of fixed cost for each participant, such as trainer/faculty transportation fees, daily subsistence allowance (DSA), and accommodation costs. In all implementation sites, the F2F component accounted for most costs, in a range between 67% and 87% of the total cost. In Nigeria and Kenya, the direct cost per participant of the BL training package were lower than the cost for delivering a full 5-day F2F training which was estimated at USD 1,144 per participant in Kenya and USD 909 in Nigeria, respectively 83% and 35% higher compared to the BL package. (Table 7).

**Table 7:** Implementation costs of the 5-day face-to-face training package in Kenya and Nigeria (USD)

|  | **Kenya** | **Nigeria** |
| --- | --- | --- |
| **No. of Participants** | 30 | 30 |
| **Cost for the implementer of the 5-day F2F training package** |  |  |
| **Resource category** | **Total Cost**<br>**(Cost per person)** | **Total Cost**<br>**(Cost per person)** |
| Staff cost during face-to-face sessions | 6,854 (228) | 4,585 (153) |
| **Total labor costs** | **6,854 (228)** | **4,585 (153)** |
| Resource fees and honoraria | 1,198 (40) | 456 (15) |
| DSA, accommodation and other allowances | 14,046 (468) | 13,856 (462) |
| Transportation fees | 2,380 (79) | 182 (6) |
| Printing, stationery costs and other consumables | 139 (5) | 2,301 (77) |
| Venues and catering for workshop/conferences | 7,469 (249) | 4,104 (137) |
| Overheads and indirect costs (7% of total cost) | 2,246 (75) | 1,784 (59) |
| **Total for the implementers of the BL training package** | **34,333 (1144)** | **27,270 (909)** |

### Strengths and Limitations of the study

This study provides insights into the implementation of a blended learning approach to in-service training in low resource settings. The feasibility study was conducted in a multi-country and multi-site context which provided opportunities for learning across the three countries as well as specific in-country experiences. The mixed methods study enhanced triangulation of findings and enabled the capturing of broad perspectives helpful in strengthening implementation of the BL approach. The self-directed learning component of the BL training is hosted on the WCEA platform which is accessible to healthcare professionals in over 40 countries in sub-Sahara Africa and accredited for continuous professional development. The BL approach was estimated to be cost saving compared to the F2F approach. Costing the BL approach increases the likelihood for adoption by policy makers.

The authors acknowledge some limitations, for example, the small sample of 89 participants which may not be generalizable to other contexts although the participants were drawn from a varied context and the sample size is within acceptable size for feasibility studies (14). This study evaluated the difference in costs between the BL and full F2F training approaches, however, it did not report on the potential differences in efficacy in reaching the intended outcomes nor the impact on the quality of care during offsite trainings. Further research is needed to understand the impact of BL trainings on quality of maternity care using the social return on investment (SROI) methodology which is recommended as a holistic approach for demonstrating value for money of interventions (23).

## Conclusion

The blended learning approach to ANC-PNC in-service training was found to be acceptable and feasible to implement in low resource settings. The BL approach was successful in improving the knowledge and skills of healthcare providers who participated in the training. Further studies are required to evaluate cost effectiveness and health provider skills of blended learning approach.

## Abbreviations

ANC: antenatal care

LMICs: low- and middle-income countries

LSTM: Liverpool School of Tropical Medicine

PNC: postnatal care

NACOSTI: National Commission for Science, Technology, and Innovation

MCQs: multiple choice questions

OSCEs: objective structured clinical examination

WCEA: World Continuing Education Alliance

## Data Availability

Data generated from the mixed methods study is included in this paper. Additional data is available from the authors upon reasonable request.

## Acknowledgments

The authors would like to thank Helen Allot for the invaluable contribution in the design of the ANC-PNC blended learning course. We would like to thank the Global Fund and Takeda Pharmaceutical Company Limited for funding the study. We are indebted to the health care professionals who participated in the ANC-PNC course and shared their experiences and ideas to strengthen future roll outs of the BL approach.

## Authors’ contributions

LAN, MF, EU, KE, MC, RM, AC, conceived and designed the study. MH, NL, MR, BR, DS, BI, OOA, KL, KE, MC, RM, EU, NF, LAN implemented the ANC-PNC Blended learning course and data collection. MC extracted and prepared data for analysis. LAN analysed qualitative data and drafted the manuscript. SW provided expertise on quantitative data and analysis. CF, AT provided expertise on costing and analysis. All authors read and approved the final manuscript before submission. AC: Funding acquisition and overall academic responsibility.

## Ethics approval and consent to participate

Ethical approval for this study was obtained from Research Ethics Committees at, Liverpool School of Tropical Medicine (ID:21-052), Nigeria (Ministry of Health Oyo State: AD13/479/44511), Kenya (Strathmore University: SU-IERC1171/21 and NACOSTI/P/21/13853), Tanzania (University of Dodoma: MA.84/261/02/‘A’/25 and Zanzibar Health Research Institute: ZAHREC/04/PR/JUNE/2022/19). A written informed consent was obtained from each participant prior to participation in the study.

## Consent for publication

Not applicable

## References

1. World Health Organization. Trends in maternal mortality 2000 to 2020: Estimates by WHO, UNICEF, UNFPA, World Bank Group and UNDESA/Population Division 2023. Available from: https://www.who.int/publications/i/item/9789240068759.

2. United Nations. Sustainable Development Goals. New York: United Nations Department of Economic and Social Affairs; 2016 23rd Nov 2019]. Available from: https://sustainabledevelopment.un.org/sdg3#targets.

3. United Nations. World Health Organization and United Nations Children’s Fund. WHO/UNICEF joint database on SDG 3.1.2 Skilled Attendance at Birth 2021. Available from: https://unstats.un.org/sdgs/indicators/database/.

4. Say L, Chou D, Gemmill A, Tunçalp Ö, Moller A-B, Daniels J, et al. Global causes of maternal death: a WHO systematic analysis. The Lancet global health. 2014;2(6):e323–e33.

5. MOH (Kenya). Saving Mothers Lives 2017, First Confidential Report into Maternal Deaths in Kenya. 2017. Available from: https://cmnh.lstmed.ac.uk/sites/default/files/content/centre-news-articles/attachments/CEMD%20Main%20Report%20Sept%203%20FINAL-%20Full%20Report.pdf.

6. WHO. Strategies towards ending preventable maternal mortality (EPMM) 2015 2nd March 2021]. Available from: https://apps.who.int/iris/handle/10665/153544.

7. Banke-Thomas A, Maua J, Madaj B, Ameh C, van den Broek N. Perspectives of stakeholders on emergency obstetric care training in Kenya: A qualitative study. International Health. 2020;12:11–8.

8. Ameh A, Mdegela M, White S, van den Broek N. The effectiveness of training in emergency obstetric care: A systematic literature review. Health Policy and Planning. 2019;34:257–70.

9. Berga K, Vadnais E, Nelson J, Johnston S, Buro K, Hu R, et al. Blended learning versus face-to-face learning in an undergraduate nursing health assessment course: A quasi-experimental study. Nurse Education Today. 2021;96(104622).

10. Yigzaw M, Tebekaw Y, Kim Y, Kols A, Ayalew F, Eyassu G. Comparing the effectiveness of a blended learning approach with a conventional learning approach for basic emergency obstetric and newborn care training in Ethiopia. Midwifery. 2019;78:42–9.

11. Shikuku DN, Tallam, E, Wako, I, Mualuko, A, Waweru, L, Nyaga, L, et al. Evaluation of capacity to deliver Emergency Obstetrics and Newborn Care updated midwifery and reproductive health training curricula in Kenya: Before and after study. 2022.

12. Shikuku DN, Tallam E, Wako I, Mualuko A, Waweru L, Nyaga L, et al. Educators’ perceptions of the early impact of COVID-19 on midwifery training in Kenya: a cross-sectional survey. International Health. 2021.

13. WHO. WHO recommendations on antenatal care for a positive pregnancy experience. Geneva, Switzerland: World Health Organization; 2016.

14. Bowen D, Kreuter M, Spring B, Cofta-Woerpel L, Linnan L, Weiner D, et al. How we design feasibility studies. American Journal of Preventive Medicine. 2009.

15. Fiorini L, Griffiths A, Houdmont J. Mixed methods research in the health sciences: A review. Malta Journal of Health Sciences. 2016.

16. Kirkpatrick D. Great Ideas Revisited: Revisiting Kirkpatrick’s Four-Level Model. Training & Development. 1996;50:54–7.

17. Teare M, Dimairo M, Shephard N, Hayman A, Whitehead A, Walters S. Sample size requirements to estimate key design parameters from external pilot randomised controlled trials: A simulation study. Trials. 2014;15:264.

18. Ladur AN, van Teijlingen E, Hundley V. ‘Whose Shoes?’ Can an educational board game engage Ugandan men in pregnancy and childbirth? BMC Pregnancy Childbirth. 2018;18(1):81.

19. Jugder N. The thematic analysis of interview data: An approach used to examine the influence of the market on curricular provision in Mongolian higher education institutions. 2016.

20. Marshall S. A Handbook for Teaching and Learning in Higher Education: Enhancing Academic Practice: Routledge; 2020.

21. Liu Q, Peng W, Zhang F, Hu R, Li Y, Yan W. The effectiveness of blended learning in health professions: Systematic review and meta-analysis. Journal of Medical Internet Research. 2016;18(1).

22. Sung Y, Kwon I, Ryu E. Blended learning on medication administration for new nurses: Integration of e-learning and face-to-face instruction in the classroom. Nurse Education Today. 2008;28:943–52.

23 Banke-Thomas, OA, Madaj, B, Ameh, C, van den Broek, N. Social return on investment methodology to account for value for money of public health interventions: A systematic review. BMC Public Health. 2015; 15:582.

